# Prediction models for adverse drug reactions during tuberculosis treatment in Brazil

**DOI:** 10.1101/2023.08.28.23294721

**Authors:** Felipe Ridolfi, Gustavo Amorim, Lauren S. Peetluk, David W. Haas, Cody Staats, Mariana Araújo-Pereira, Marcelo Cordeiro-Santos, Afrânio L. Kritski, Marina C. Figueiredo, Bruno B. Andrade, Valeria C. Rolla, Timothy R. Sterling, Regional Prospective Observational Research in Tuberculosis (RePORT)-Brazil consortium

## Abstract

**Background:** Tuberculosis (TB) treatment-related adverse drug reactions (TB-ADR) can negatively affect adherence and treatment success rates.

**Methods:** We developed two prediction models for TB-ADR. We included drug-susceptible pulmonary TB participants who initiated standard TB therapy. TB-ADR were determined by physician-assigned attributions of causality, and described according to affected organ system, timing, and grade. Potential predictors of TB-ADR included concomitant medication (CM) use, HIV-status, glycated hemoglobin (HbA1c), age, body mass index (BMI), sex, substance use, and TB drug metabolism variables (e.g., *NAT2* acetylator profile). Bootstrapped backwards selection was used to develop the models. Cox proportional hazards regression was used to evaluate TB-ADR risk.

**Results:** There were 156 TB-ADR among 102 (11%) of the 945 participants included. Most TB-ADR were hepatic (n=82;53%), grade 2 (n=121;78%), and occurred in *NAT2* slow acetylators (n=62;61%). The main prediction model included CM use, HbA1c, alcohol-use, HIV-infection, BMI, and age. The alternative model included the same variables, except replaced BMI with *NAT2*. Both models had good performance and fit. CM use and HIV-infection increased TB-ADR risk.

**Conclusions:** The model with only clinical variables and that with *NAT2* were highly predictive of TB-ADR. The *NAT2* model provides rationale to evaluate isoniazid dose adjustment and ADR risk.

## Introduction

First-line tuberculosis (TB) treatment is effective, resulting in high cure rates for drug-susceptible disease when the standard 6-month regimen is completed.^1–3^ However, adverse drug reactions (ADR) influence rates of treatment completion and effectiveness, through interrupted therapy and the need to use alternative regimens.^4^ Recent data from a prospective cohort study in Brazil found that more than three-quarters of participants experienced at least one ADR episode during TB treatment; over half of all participants had clinical symptoms of ADR, whereas the remainder were diagnosed based on laboratory measurements.^5^ Moreover, treatment interruptions due to ADR occur in up to 15% of patients, and often within the initial two-month intensive phase of treatment.^5,6^ This can lead to treatment modification, requiring longer and less effective regimens, and drugs that are more expensive and more toxic. Even when ADR do not result in treatment modification, they can potentially affect adherence and negatively impact treatment success rates.^7^

Our group previously developed a prediction model for unsuccessful TB treatment outcomes, including death, treatment failure, regimen switch, and incomplete treatment.^8^ However, the model did not evaluate ADR, which is a measure of drug safety rather than effectiveness. Identifying persons at increased risk of TB-related ADR (TB-ADR), especially modifiable risk factors, could facilitate interventions to lower the risk of ADRs. Given the importance of TB drug tolerability for treatment completion and effectiveness, and the lack of models to predict TB treatment-related ADR, we developed prediction models for ADR during TB treatment in a large, prospective observational cohort in Brazil.^9^ We developed two models: one with only clinical variables, and one that also included information on TB drug metabolizer polymorphisms.

## Methods

### Study design and population

Our study included participants enrolled in the Regional Prospective Observational Research in Tuberculosis (RePORT) Brazil cohort. RePORT-Brazil is a prospective observational cohort study of persons with newly-diagnosed, culture-confirmed, pulmonary TB at five sites across three regions in Brazil, enrolled between June 2015 and June 2019. Participants were followed for two years.^9,10^ RePORT-Brazil excluded participants who had previously received anti-TB therapy for ≥7 days, received >7 days of fluoroquinolone therapy within 30 days of enrollment, were pregnant or breastfeeding, or did not plan to remain in the enrollment region during follow-up. Sites were in Rio de Janeiro (Instituto Nacional de Infectologia Evandro Chagas, Clínica de Saúde Rinaldo Delmare, Secretaria de Saúde de Duque de Caxias), Salvador (Instituto Brasileiro para Investigação da Tuberculose), and Manaus (Fundação Medicina Tropical Dr. Heitor Vieira Dourado). The study population has been shown to be representative of all TB cases reported to the national Brazilian TB registry (SINAN).^9^

For this study, we included RePORT-Brazil participants with drug-susceptible pulmonary TB who initiated standard TB therapy, comprising a two-month intensive phase of isoniazid, rifampicin or rifabutin, pyrazinamide, and ethambutol, followed by a four-month continuation phase of isoniazid with either rifampicin or rifabutin, all dosed according to the participant’s weight.^11,12^

### Data collection and definitions

Clinical, demographic, and outcome data were collected longitudinally at in-person study visits at baseline, month 1, month 2, and end of treatment (typically month 6), and then via telephone follow-up every six months until month 24. Data were stored in REDCap.^13^

RePORT-Brazil used a symptom-based approach to identify ADR during TB treatment. This analysis included any TB-ADR based on physician-assigned attribution of “possibly”, “likely”, or “definitely” related to TB treatment.^14^ TB-ADR were also described according to the affected organ system (hepatic, dermatologic, neurologic), and by grade (Grade 2, 3, 4, and 5).^11,15^ Grade 2 reactions were considered moderate severity, Grade 3 were severe, Grade 4 were life-threatening, and Grade 5 were associated with death. Grade 1 reactions were not captured in RePORT-Brazil. Outcome definitions followed the recently-updated World Health Organization TB treatment outcome definitions^16^: cure, treatment completion, treatment failure, death from any cause during treatment, and loss to follow-up (LTFU). Treatment failure is now defined as participants whose TB regimen needed to be terminated or permanently changed to a new regimen or treatment strategy due to lack of clinical and/or bacteriological response, ADR that resulted in treatment discontinuation, or evidence of acquisition of drug resistance. LTFU included treatment abandonment, lost contact, patient transfer, participant withdrawal, and not evaluated.

All participants underwent HIV testing at baseline unless already known to be persons with HIV (PWH), and each drug that comprised the antiretroviral regimens (usually a 3-drug combination) was counted separately as a CM in the group of medications for chronic disease, as mentioned above. Among PWH, we assessed baseline CD4 T-cell count and HIV-1 RNA viral load (VL), and both variables were evaluated as either continuous or categorical variables; we considered a breakpoint of 200 cells/mL for categorical CD4 T-cell count, and for VL a cutoff of <1000 copies/mL to define virologic suppression. We also considered timing of antiretroviral therapy (ART) initiation. Prior ART use was defined as exposure to any ART drug before TB diagnosis, ART-naïve was defined as having never received ART before TB diagnosis.

Diabetes status at baseline was based on both self-reported history of diabetes mellitus and glycosylated hemoglobin (HbA1c) levels. HbA1c <5.7% was considered no diabetes, HbA1c ≥5.7% to <6.5% was considered pre-diabetes, and HbA1c ≥6.5% was considered diabetes.^17,18^ Age was considered in years at the time of enrollment, and BMI was categorized as underweight (BMI ≤18.5 kg/m^2^), normal (>18.5 and ≤25 kg/m^2^), and overweight (>25 kg/m^2^).^19^ Alcohol, tobacco, and drug use were self-reported and categorized as never, former or current use. Additional data, such as smear positivity at baseline, presence of cavitation on chest x-ray, and other concomitant chronic diseases were also collected. The variable race was self-reported at baseline.

We also evaluated concomitant medication (CM) use (other than anti-TB treatment) at baseline, classified by mechanism of action (e.g., anti-bacterial, oral hypoglycemic, analgesic, corticosteroids) and considering the number of CM used, which were grouped into four categories: 1) medications for chronic disease treatment (e.g., hypertension, diabetes, HIV); 2) pain/allergy medications, such as analgesic/antipyretic, antihistaminic, corticosteroid, and non-steroidal anti-inflammatory; 3) antimicrobial medications, including antibiotics, antifungals, and antivirals other than antiretrovirals; and 4) miscellaneous medications, including antiemetic, vitamins, supplements, and any other medications not previously classified.

Genotyping was done of selected single nucleotide polymorphisms (SNPs) in genes relevant to metabolism of TB drugs. Genotyping was done by VANTAGE (Vanderbilt Technology for Advanced Genomics) using MassARRAY® iPLEX Gold (Agena Bioscience™, California, USA) and Taqman (ThermoFisher Scientific, Massachusetts, USA). We genotyped 4 SNPs to categorize *NAT2* acetylator groups, which have been associated with higher INH concentrations and higher incidence of TB-ADR including hepatotoxicity^20–22^, as slow, intermediate, or rapid acetylators.

We selected the candidate predictors for the two prediction models incorporating information from a comprehensive literature review and biological plausibility. ^20–25^ Considerable deliberation was given to the inclusion of the following variables: CM use, HIV status (positive or negative), HbA1c, age, body mass index (BMI), alcohol use, tobacco use, drug use, sex, and the genetic data.

### Statistical analysis

Participant characteristics were described stratifying by TB-ADR occurrence (any TB-ADR and no TB-ADR). Details of each ADR were described, including the grade, physician-assigned attribution of relation to TB treatment, type of ADR (organ system affected), and timing of ADR. For all descriptive analyses, continuous variables were summarized with median and interquartile range (IQR) and categorical variables by frequency and percentages.

We used two approaches for the models to predict risk of any TB-ADR. First, we considered the applicability and utility of a prediction score in daily practice, and included nine pre-specified clinical and sociodemographic predictors to build the main model. Secondly, since we also had data on *NAT2* acetylator group, we built an alternative model, with twelve pre-specified predictors. The methods used in prediction model development have been described in detail elsewhere.^8,26^ Bootstrapped backwards selection was used for models selection.^27^ Performance was evaluated with discrimination and calibration measures to assess the accuracy and reliability of the prediction model. Discrimination was quantified with the c-statistic.^28^ Calibration was assessed using calibration plot, calibration intercept, and calibration slope^29^ (**Figure 1**).

**Figure 1.**
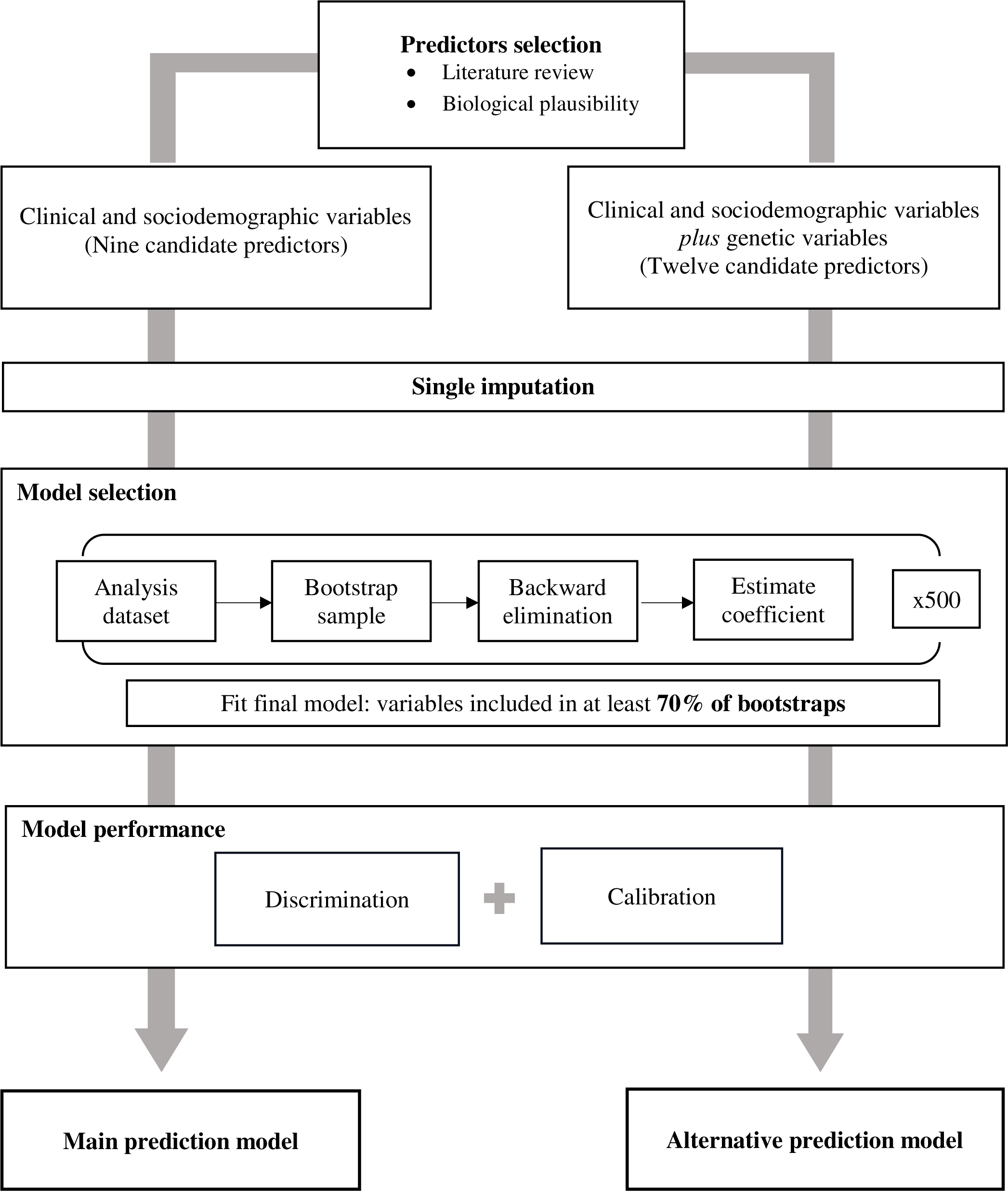
Flow chart with the steps followed to build the prediction models **Legend**. Schematic representation of each model development step and assessment of model performance. **Predictors selection**. A comprehensive literature review, taking into account biological plausibility, was made to define which predictors to pre-select for the models. Considering the applicability in daily practice, we included nine easy-available clinical and sociodemographic predictors for the main model. For the alternative model, we decided to also include genetic information related to TB drugs metabolism, such as isoniazid and rifampicin. *NAT2* genotypes, related to isoniazid metabolism, were categorized based on combinations of gene variants rs1801280 (*NAT2*\*5), rs1799930 (*NAT2*\*6), rs1799931 (*NAT2*\*7), and rs1801279 (*NAT2*\*14). The slow acetylator was defined as homozygous for the variant allele at any locus or heterozygous at 2 or more loci; intermediate as heterozygous at a single locus; or rapid if no variant allele. And two SNPs related to hepatotoxicity and the metabolism of rifampicin, rs11045819 (gene *SLCO1B1*) and rs412543 (gene *GSTM1*) were also genotyped. **Single imputation** was used as fewer than 1% of participants had any missing data; all continuous variables were modeled with restricted cubic splines with 3 knots to account for non-linear relationships. **Bootstrapped backwards selection** was used for model selection. This approach involved repeated iterations of backwards selection in bootstrap samples (with replacement) from the data to evaluate the importance of each candidate predictor. After 500 iterations of bootstrapped backwards selection, variables that were selected in at least 70% of bootstrap samples were included in the final model. **Model performance** was evaluated with discrimination and calibration measures to assess the accuracy and reliability of the prediction model. Discrimination was quantified with the c-statistic. Calibration was assessed using calibration plot, calibration intercept, and calibration slope. Internal validation with bootstrap resampling was used to estimate optimism-corrected performance measures. Predictions from the final model accounted for shrinkage according to the heuristic shrinkage factor, estimated as χ^2^_model_ – df / χ^2m^^odel^, where “χ^2^_model_” is the chi-square model and “df” is degrees of freedom.

We additionally performed Cox proportional hazard regression to evaluate associations between each variable selected for the final primary prediction model and the risk of TB-ADR (but not for the alternative model). Participants could experience multiple ADR over the course of treatment, but each analysis evaluated time until the first ADR. Individuals who were LTFU were censored at their last kept study visit.^30^ For all analyses, confounders were selected *a priori* and included age, sex, and smoking and alcohol status at baseline. For the diabetes analyses, HbA1c was considered as a continuous variable (modeled with a restricted cubic spline with 3 knots) and as a categorical variable, to create diabetes categories of no diabetes, pre-diabetes, and diabetes, as described above. Fewer than 1% of participants had any missing data, so single imputation with predictive mean matching was used in the analysis. All analyses were conducted using R software (version 4.2.0).

## Results

Of the 945 participants included, 102 (11%) experienced an ADR (**Supplemental Figure 1**). Among those who experienced ADRs, most were men (68%, n=69), 37% (n=38) were PWH, 80% (n=82) had CM use at baseline, and most were slow *NAT2* acetylators (61%, n=62) (**Table 1)**. Overall, there were 156 TB-ADR episodes reported; most of TB-ADR occurred during the intensive phase of TB treatment (N=120, 77%). Most TB-ADR were Grade 2 (n=121, 78%), and the most frequent type was hepatic (n=82, 53%), followed by dermatologic (n=35, 22%), and neurologic (N=17, 11%) (**Table 2**).

**Table 1.** Baseline clinical and demographic characteristics of the study population (N=945).

| Characteristics | Any TB-ADR<br>(N=102) | No TB-ADR<br>(N=843) |
| --- | --- | --- |
| N (%) or Median [IQR] |  |  |
| Age | 38 (26 – 49) | 35 (25 – 49) |
| Female sex | 33 (32) | 284 (34) |
| Self-reported race |  |  |
| Brown | 13 (13) | 228 (27) |
| Black | 60 (59) | 431 (51) |
| White | 0 (0) | 20 (2.3) |
| Other/Unknown | 29 (28) | 164 (19) |
| NAT2 acetylator group<sup>1</sup> |  |  |
| Rapid | 5 (5) | 73 (9) |
| Intermediate | 35 (34) | 348 (41) |
| Slow | 62 (61) | 422 (50) |
| BMI at baseline, kg/m<sup>2</sup> | 20.6 (19.1 – 22.2) | 20.1 (18.2 – 22.5) |
| BMI category |  |  |
| Underweight (<18.5) | 72 (71) | 517 (61) |
| Normal (18.5 – 25) | 11 (11) | 82 (9.7) |
| Overweight (>25) | 19 (19) | 244 (29) |
| Sputum smear positive for acid-fast bacilli | 76 (75) | 694 (82) |
| Cavitation on chest x-ray | 30 (31) | 439 (52) |
| History of diabetes (self-report) | 6 (5.9) | 105 (12) |
| HbA1c at baseline | 5.8 (5.5–6.2) | 5.9 (5.4–6.4) |
| Diabetes (HbA1c category) | 6 (5.9) | 105 (12) |
| Normal (<5.7) | 35 (35) | 321 (38) |
| Pre-diabetes (5.7 – 6.5) | 47 (47) | 312 (37) |
| Diabetes (≥6.5) | 19 (19) | 204 (24) |
| PWH | 38 (37) | 145 (17) |
| Other chronic disease<sup>2</sup> | 23 (23) | 193 (23) |
| Receiving concomitant medication at baseline | 82 (80) | 336 (40) |
| Alcohol use |  |  |
| Never | 17 (17) | 135 (16) |
| Former | 56 (55) | 310 (37) |
| Current | 29 (28) | 398 (47) |
| Tobacco use |  |  |
| Never | 50 (49) | 406 (48) |
| Former | 32 (31) | 241 (29) |
| Current | 20 (20) | 196 (23) |
| N (%) or Median [IQR] |  |  |
| Drug use<sup>3</sup> |  |  |
| Never | 69 (68) | 555 (66) |
| Former | 26 (25) | 177 (21) |
| Current | 7 (6.9) | 110 (13) |

**Abbreviations:** TB-ADR: TB treatment-related adverse drug reaction; IQR: interquartile range; BMI: body mass index; PWH: people with HIV;
<sup>1</sup> NAT2 acetylator group is defined by 4 single gene polymorphisms.
<sup>2</sup> Other chronic disease includes: cardiovascular disorders (e.g., hypertension, coronary artery disease, peripheral artery disease, post-stroke disorders), thyroid disorders (e.g., hypo- and hyperthyroidism), pulmonary disorders (e.g., asthma, chronic obstructive pulmonary disease), psychiatric disorders (e.g., depression), and chronic kidney disorders.
<sup>3</sup> Drug use includes all drugs, such as marijuana, cocaine, crack, ecstasy, injectable drugs, inhaled solvents, oxycodone, and cocaine paste base.

**Table 2.** Distribution of all TB treatment-related adverse drug reactions (event-level) among 102 persons (11%) with a TB treatment-related adverse drug reaction.

|  |  | **N=156**<br>**N (%)** |
| --- | --- | --- |
| **Grade** |  |  |
|  | Grade 2 | 121 (78) |
|  | Grade 3 | 25 (16) |
|  | Grade 4 | 9 (5.8) |
|  | Grade 5 | 1 (0.6) |
| **Relation to TB treatment<sup>1</sup>** |  |  |
|  | Related | 21 (13) |
|  | Likely | 71 (46) |
|  | Possible | 64 (41) |
| **Type of TB-ADR** |  |  |
|  | Hepatic | 82 (53) |
|  | Dermatologic | 35 (22) |
|  | Neurologic | 17 (11) |
|  | Other | 22 (14) |
| **Timing of ADR in relation to TB treatment start** |  |  |
|  | Within 1 month | 79 (51) |
|  | 1-2 months | 41 (26) |
|  | 2-3 months | 15 (9.7) |
|  | 3-4 months | 5 (3.2) |
|  | 4-5 months | 10 (6.5) |
|  | 5-6 months | 4 (2.6) |
|  | ≥6 months | 1 (0.6) |

**Footnotes:** Neurologic is composed of 13 events of peripheral neuropathy and 4 other neurologic events. Other ADR include: anemia (n=2), arthralgia (n=5), cardiovascular/respiratory (n=3), musculoskeletal (n=2), reproductive (n=2), sensory (n=1), uric acid elevation (n=6), and urinary/renal (n=1).
<sup>1</sup> This classification establishes the causality of the adverse drug-reaction and is defined based on Naranjo et.al: “A ‘related’ reaction followed a reasonable temporal sequence after a drug or in which a toxic drug level had been established, followed a recognized response to the suspected drug, and was confirmed by improvement on withdrawing the drug and reappeared on re-exposure. A ‘likely’ reaction has a reasonable temporal sequence after a drug, followed a recognized response to the suspected drug, was confirmed by withdrawal but not by exposure to the drug, and could not be reasonably explained by the known characteristics of the patient's clinical state. And a ‘possible’ reaction followed a temporal sequence after a drug, possibly followed a recognized pattern to the suspected drug, and could be explained by characteristics of the patient's disease.” (Naranjo CA, Busto U, Sellers EM, Sandor P, Ruiz I, Roberts EA, et al. A method for estimating the probability of adverse drug reactions. Clin Pharmacol Ther. 1981 Aug;30(2):239–45)

After bootstrapped backward selection, six variables were most predictive of TB-ADR and were included in the main model: CM use at baseline, HbA1c, alcohol use, HIV status, BMI, and age (**Table 3**). The model with these variables, including restricted cubic splines with three knots for HbA1c and age, demonstrated good performance with a c-statistic of 0.79 (95%CI: 0.75-0.83) and the calibration curve indicated a good fit, with an optimism-correct intercept and slope of -0.22 and 0.87, respectively. A shrinkage factor of 0.90 was applied to correct for uncertainties introduced in model development and improve fit in external validation (**Figure 2**). The alternative prediction model included the following variables: CM use, HbA1c, alcohol use, *NAT2* acetylator group, age, and HIV status. This model also demonstrated good performance, with a c-statistic of 0.79 (95%CI: 0.74-0.83). The calibration curve indicated a good fit, with an optimism-correct intercept and slope of -0.09 and 0.94, respectively (**Supplemental Table 1 and Supplemental Figure 2)**. The predicted risks from the final model can be applied to new populations using a nomogram (**Supplemental Figure 3**).

**Figure 2.**
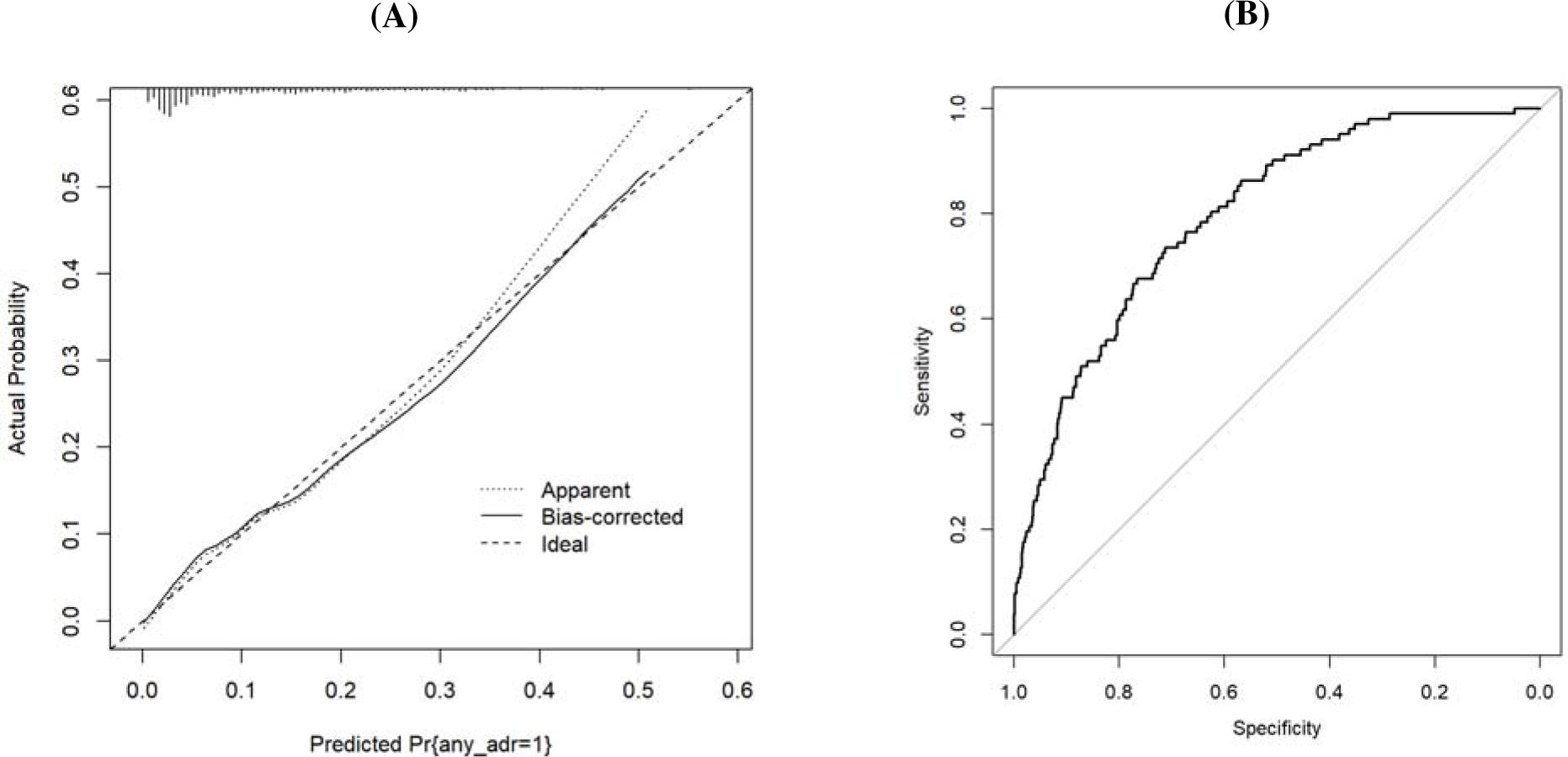
Performance of the main prognostic model **Footnote: (A)** The calibration plot displays agreement between observed and predicted outcome probabilities across deciles of outcome (TB-treatment related adverse drug reaction (TB-ADR)) risk. An ideal calibration curve has an intercept of 0 and a slope of 1 (dashed line). The apparent calibration (dotted line) is calibration of the model in the original data, and the bias-corrected line is corrected for overfitting using 500 bootstrap samples. The bias-corrected calibration intercept and slope were -0.22 and 0.87, respectively. The top of the plot displays a histogram of the distribution of predicted probabilities TB-ADR for the 945 culture-confirmed, drug-susceptible pulmonary tuberculosis participants included in the study. A shrinkage factor of 0.90 was applied to correct uncertainties introduced in model development and improve fit in external validation. **(B)** The receiver operating characteristic (ROC) curve measures discrimination of the model, i.e., how well the model can differentiate between those with and without an outcome. The area under the ROC curve, which is equivalent to the c-statistic, is 0.79 (95% CI: 0.75-0.83).

**Table 3.**
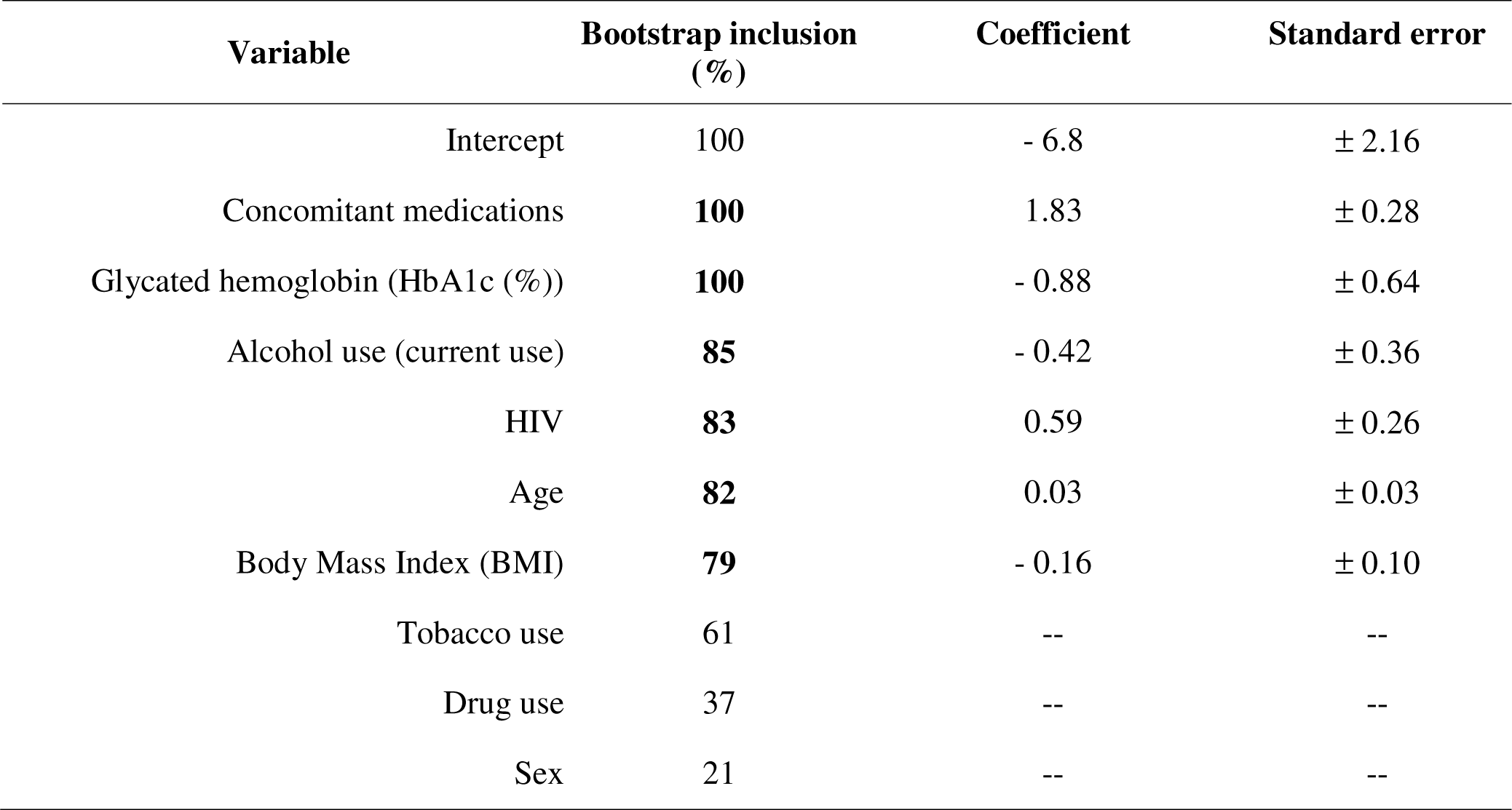
Main prediction model for any TB treatment-related adverse drug reaction.

| **Variable** | **Bootstrap inclusion (%)** | **Coefficient** | **Standard error** |
| --- | --- | --- | --- |
| Intercept | 100 | - 6.8 | ± 2.16 |
| Concomitant medications | **100** | 1.83 | ± 0.28 |
| Glycated hemoglobin (HbA1c (%)) | **100** | - 0.88 | ± 0.64 |
| Alcohol use (current use) | **85** | - 0.42 | ± 0.36 |
| HIV | **83** | 0.59 | ± 0.26 |
| Age | **82** | 0.03 | ± 0.03 |
| Body Mass Index (BMI) | **79** | - 0.16 | ± 0.10 |
| Tobacco use | 61 | -- | -- |
| Drug use | 37 | -- | -- |
| Sex | 21 | -- | -- |

In addition to the prediction model, we evaluated the association of each predictor included in the main model, individually, with TB-ADR occurrence. CM use at baseline increased by ∼5 times the risk of any TB-ADR (hazard ratio (HR) 5.38, 95% confidence interval (CI): 3.25-8.89). PWH also had an increased risk of any TB-ADR (HR 2.68, 95%CI 1.75-4.09) and of hepatic TB-ADR (HR 5.26, 95%CI 2.63-10.52, graph not shown). Higher levels of HbA1c were associated with decreased the risk of having an TB-ADR (**Figure 3**). For example, compared to individuals with HbA1c=5.5%, individuals with an HbA1c=9% had 63% decreased risk of TB-ADR (HR 0.47; 95%CI: 0.23-0.95) and individuals with HbA1c of 11% had 83% decreased risk of TB-ADR (HR 0.17; 95%CI: 0.04-0.66) (**Supplemental Figure 4**).

**Figure 3.**
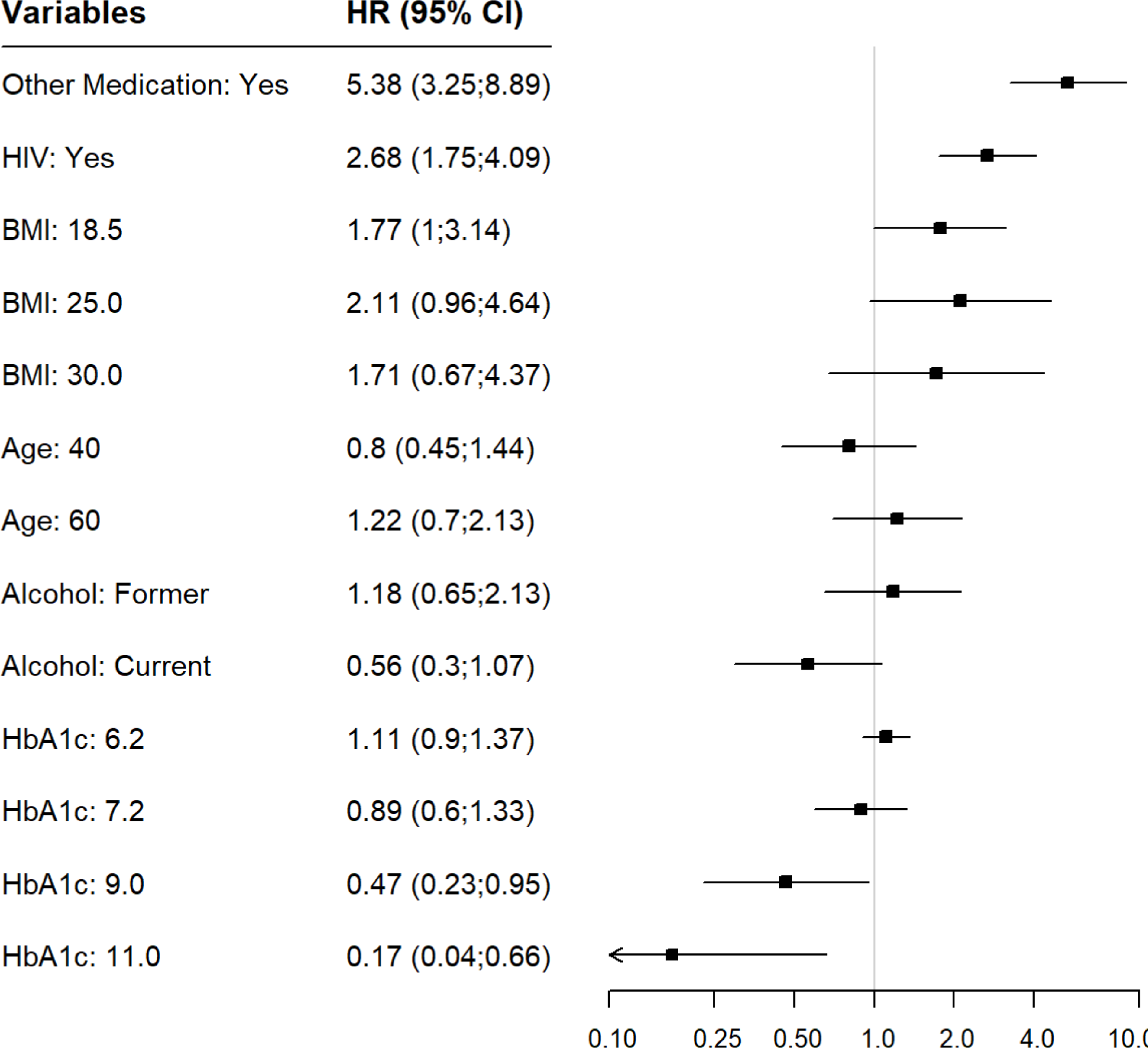
Coefficient plot of variables and associations with TB treatment-related adverse drug reaction. **Footnote**: HR; hazard ratio. 95% CI: 95% confidence interval. Reference categories for other medication: no; for HIV: no/HIV-negative; Body Mass Index (BMI): 15, for age: 20; for alcohol: never; for HbA1c: 5.5.

Since CM use and HIV status were highly associated with TB-ADR, we performed additional sub-analysis to assess the association of CM and TB-ADR, stratifying by HIV status. A subset of patients (n = 315) also had information about the type and number of medications used at baseline, allowing us to assess the effect of polypharmacy on the outcome instead of a simple binary (yes/no) exposure. HIV-seronegative participants who were prescribed at least one CM at baseline had a higher risk of having a TB-ADR compared to participants with no CM (adjusted HR (aHR) 6.89, 95%CI: 3.63-13.07). As the number of CM increased among HIV-negative participants, no substantial increase in the TB-ADR risk was observed. Among PWH, no association with number of CM and TB-ADR was found (**Figure 4**). Regarding the type of CM, we categorized 27 classes of CM into 4 groups. Among participants taking 4 or more CM medications for chronic diseases and antimicrobials were more frequent. Most participants taking only one CM were using pain medication (70.6%), and within this group, only 12% (N=13) were acetaminophen-containing drugs (**Table 4)**.

**Figure 4.**
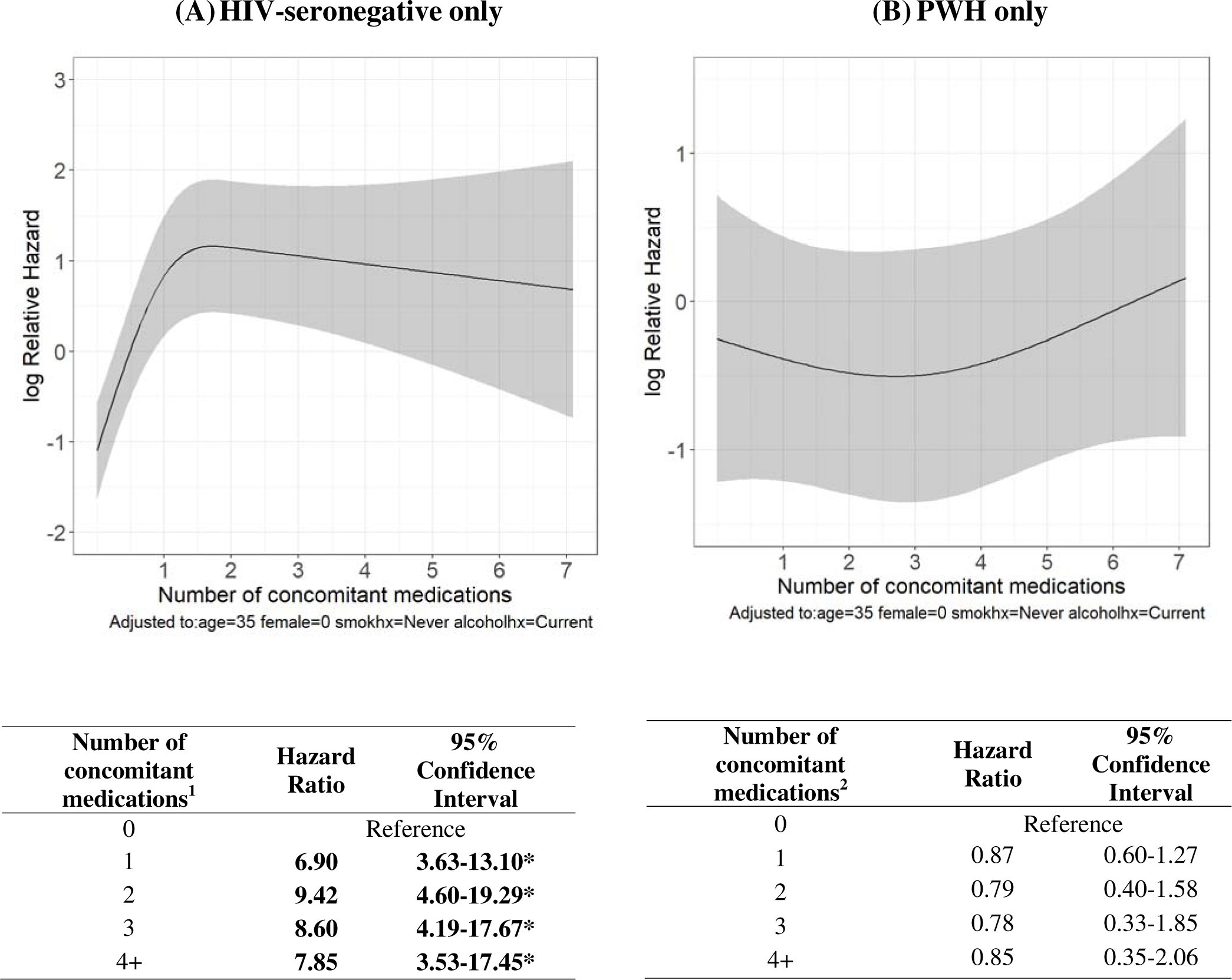
Association between concomitant medication and TB-ADR, stratified by HIV status. <u>Abbreviations</u>: TB-ADR: TB treatment-related adverse drug reaction; PWH: People with HIV. **Footnote**: (A) Risk of TB-ADR among participants HIV-seronegative, according to the number of concomitant medications. ^1^ Number of participants in each subgroup: for 0, N=469; for 1, N=129; for 2, N=49, for 3, N=13, for 4+, N=19. **\*** The risk of TB-ADR increases the highest from 0 to 1 concomitant medication. (B) Among participants living with HIV, no risk of TB-ADR was observed according to the number of concomitant medications. ^2^ Number of participants in each subgroup: for 0, N=40; for 1, N=14; for 2, N=15, for 3, N=25, for 4+, N=69.

**Table 4.** Number of concomitant medications according to medication class/mechanism of action.

| Medication groups/classes N (%) | Number of concomitant medications<sup>1</sup> |  |  |  |
| --- | --- | --- | --- | --- |
|  | 1<br>(N=143) | 2<br>(N=64) | 3<br>(N=38) | 4+<br>(N=88) |
| Chronic diseases | 21 (13.7) | 35 (25.4) | 40 (38.1) | 96 (34.4) |
| Pain/allergy medication | **108 (70.6)** | 69 (50) | 22 (21) | 54 (19.4) |
| Acetaminophen containing: No | 95 (88) | 65 (94.2) | 22 (100) | 50 (92.6) |
| Acetaminophen containing: Yes | 13 (12) | 4 (5.8) | - - | 4 (7.4) |
| Antimicrobial | 13 (8.5) | 23 (16.7) | 30 (28.6) | 96 (34.4) |
| Miscellaneous | 11 (7.2) | 11 (8) | 13 (12.4) | 33 (11.8) |

**Footnote:** Chronic disease medications included anti-hypertensives, oral hypoglycemics, anti-depressants. Pain/allergy medications included analgesic/antipyretic, non-steroidal anti-inflammatory, corticosteroids, and anti-histaminic. Antimicrobials included antivirals other than antiretrovirals, antibiotics, and antifungals. Miscellaneous included vitamins, supplements, and all other medications that were not classified in previous three groups. <sup>1</sup> The subgroups are not mutually exclusive meaning that one participant can be counted.

Further analysis considering HIV-related variables found no association with TB-ADR when comparing CD4 T-cell counts (>200 vs <200), VL (suppressed vs non-suppressed) or ART exposure (ART-experienced vs ART-naïve) (**Supplemental Figure 5**)

## Discussion

This analysis highlights several important findings. Most TB-ADR were of moderate severity (Grade 2), were hepatic, and occurred in the intensive phase of TB treatment. Use of concomitant medication at baseline, HbA1c, alcohol use, HIV status, BMI, and age were predictive of TB-ADR. Our findings of ADR severity, of organ system affected, and of the timing of ADR during TB treatment are consistent with previous studies.^5,23^

The two models developed were highly predictive of TB-ADR. Interestingly, most of the predictors were common to both models. The main model included variables that can be easily obtained in the clinical setting, and can promptly demonstrate the risk of TB-ADR in a person starting TB treatment. The alternative model included the same variables plus the *NAT2* acetylator profiles, which is generally not readily available in clinical practice. However, such information may be useful for evaluating interventions that could improve the tolerability of TB treatment, such as isoniazid dose adjustment.

When evaluating associations between predictors and risk of TB-ADR, CM use at baseline and HIV coinfection were associated with increased the risk for TB-ADR, while high levels of HbA1c were associated with decreased risk for TB-ADR. Among HIV-seronegative participants, CM use at baseline was strongly associated with risk of TB-ADR, especially when comparing no CM versus one CM. The most common class of CM used by participants taking only one CM was non-acetaminophen-containing pain medication. In Brazil, dipyrone (i.e., Metamizole) is a very commonly used analgesic and antipyretic drug, and is available without prescription. Dipyrone has a potential hepatotoxic effect; however, the most well-known ADR for this drug is agranulocytosis – which occurs infrequently and was not observed in our study.^31,32^ Among PWH, there was no significant association between CM use, which included ART, and the risk of TB-ADR. There are limited previous studies evaluating this association, however, the available literature is not in accordance with our findings. A previous study demonstrated increased hepatotoxicity risk with concomitant anti-TB-HIV therapy, and that ART alone was associated with 3 times the risk of ADR compared to TB treatment alone.^33^ Moreover, a recent study found higher incident rates of liver injury among TB-HIV patients, especially when starting ART during the intensive phase of TB treatment.^34^

In the present analysis, PWH were at increased risk for TB-ADR, including hepatic ADR, which is consistent with several previous studies,^34,35^ and therefore an expected finding. However, we did not find an association between HIV-related factors, such as CD4 T-cell count or VL, and TB-ADR in our study.

Interestingly, the risk of TB-ADR decreased as baseline HbA1c increased – i.e., participants with higher HbA1c levels had lower risk of TB-ADR. Although the reason for this association is unclear, it could be due to decreased drug absorption among persons with poor diabetes control, and therefore lower TB drug exposures.^36,37^ The association between BMI, age, and alcohol use were inconclusive for ADR risk. Moreover, there was a higher frequency of TB-ADR among slow *NAT2* acetylators, which is in accordance with the literature ^21,22,38^

There were several limitations of this study. We considered only the first TB-ADR occurrence; TB-ADR can overlap or occur more than once during TB treatment. In addition, the determination of attribution of ADRs to TB drugs by local care providers in this open-label study could be biased. Due to the high number of classes of CM received by the study population (i.e., 27), we could not assess the association between any specific class (e.g., antiretroviral, anti-hypertensives) and ADR risk with TB treatment. We considered only baseline information regarding CM use, and some participants used multiple classes of medication simultaneously.

Depending on the type of medication, it may be taken regularly (e.g., for chronic diseases), or intermittently (e.g., for symptom relief). There was a relatively low rate of TB-ADR (11%) compared to rates in the literature^39,40^ and this may have affected the performance of the prediction model. Moreover, the lack of association between CM and risk of TB-ADR among PWH may have been due to the sample size; and because we only considered baseline information about ART, and not the possible time-varying association of ART and TB-ADR. We included only participants with culture-confirmed, drug-susceptible, pulmonary TB on standard therapy regimens for 6 months, which could affect the generalizability of the results.

This study had several strengths. This was a large multicenter prospective cohort study, representative of persons with TB in Brazil^9^, with uniform data collection and regularly scheduled visits. Additionally, the variables included in the model are readily available in clinical settings, which, in turn, can make implementation of the prediction model feasible. We also developed an alternative model which included genetic information. We are not aware of previous studies that developed a prediction model for ADR during TB treatment.

In conclusion, we developed two models that were highly predictive of TB treatment-related ADR; one with easy-to-assess variables, and another which included genetic data. Additionally, we evaluated associations between important risk factors and TB-ADR. Knowledge of these factors at the time of TB treatment initiation, and interventions to decrease their contribution (e.g., fewer concomitant medications or isoniazid dose adjustment) could improve TB treatment tolerability.

## Data Availability

All data produced in the present study are available upon reasonable request to the authors

## Author contributions

F.R., G.A., and L.S.P. conceptualized the research question and drafted the initial manuscript. L.S.P. and G.A. conducted the analysis. V.R. and T.R.S. provided thorough feedback on the research design and analysis interpretation, supervised the analysis, and revised successive drafts of the manuscript. B.A., M.A.P., and D.W.H. provided valuable feedback and comments on successive manuscript drafts. B.A., M.C.S., M.T., A.K., T.R.S., V.R., and M.C.F. played pivotal roles in the conceptualization of the RePORT-Brazil cohort, project administration, data and funding acquisition, and revised successive drafts of the manuscript. All authors approved the final version of the manuscript.

## Ethical approval

All clinical investigations were conducted according to the principles of the Declaration of Helsinki. The RePORT-Brazil protocol, informed consent, and study documents were approved by the institutional review boards at each study site and at Vanderbilt University Medical Center. Participation in RePORT-Brazil was voluntary, and written informed consent was obtained from all such participants.

## Acknowledgements

The authors thank the study participants, the teams of clinical and laboratory platforms of all RePORT Brazil consortium sites.

