## Supplemental Material for "Prediction models for adverse drug reactions during tuberculosis treatment in Brazil"

**SUPPLEMENTARY MATERIAL**

Culture-confirmed, Pulmonary TB

RePORT Brazil Study participants

N=1,076

Participants **included**

(Study population)

N=945

**Exclusions** (N= 131)

82 (62%) Drug-resistant cases

8 (6%) Non-standard TB treatment

41 (32%) Extrapulmonary TB cases

requiring extended treatment duration

Bone/joint (2, 5%)

Cutaneous (1, 2.5%)

Disseminated (7, 17%)

Intestinal (8, 19%)

Laryngeal (16, 39%)

Meningeal (6, 15%)

Spleen (1, 2.5%)

Participants with

**any TB treatment-related** **adverse drug reaction,**

≥ **Grade 2**

N=102 (11%)

Participants **without TB treatment-related**

adverse drug reaction

N=843

**Supplemental Figure 1**. Study diagram

**Supplemental Table 1**. Alternative prediction model, with genetic variables, for any TB treatment-related adverse drug reaction.

| **Variable** | **Bootstrap inclusion**  **(%)** | **Coefficient** | **Standard error** |
| --- | --- | --- | --- |
| Intercept | **100** | - 2.87 | ± 0.69 |
| Concomitant medications | **100** | 1.82 | ± 0.27 |
| Glycated hemoglobin (HbA1c (%)) | **100** | - 0.30 | ± 0.09 |
| Alcohol use (current use) | **91** | - 0.45 | ± 0.33 |
| *NAT2* acetylator profile^1^ | **79** | 0.35 | ± 0.18 |
| HIV | **77** | 0.49 | ± 0.25 |
| Age | **76** | 0.02 | ± 0.01 |
| Body Mass Index (BMI) | 62 | -- | -- |
| Tobacco use | 56 | -- | -- |
| Drug use | 45 | -- | -- |
| rs11045819 (*SLCO1B1*) | 40 | -- | -- |
| rs412543 (*GSMT2*) | 20 | -- | -- |
| Sex | 16 | -- | -- |

**Footnote**:

^1^*NAT2* acetylator profile included rapid, intermediate, and slow groups; The interpretation of this variable within the model was that as the *NAT2* acetylator slowed, there was an increased probability of TB-ADR to occur.

**Supplemental Figure 2.** Performance of the alternative prognostic model

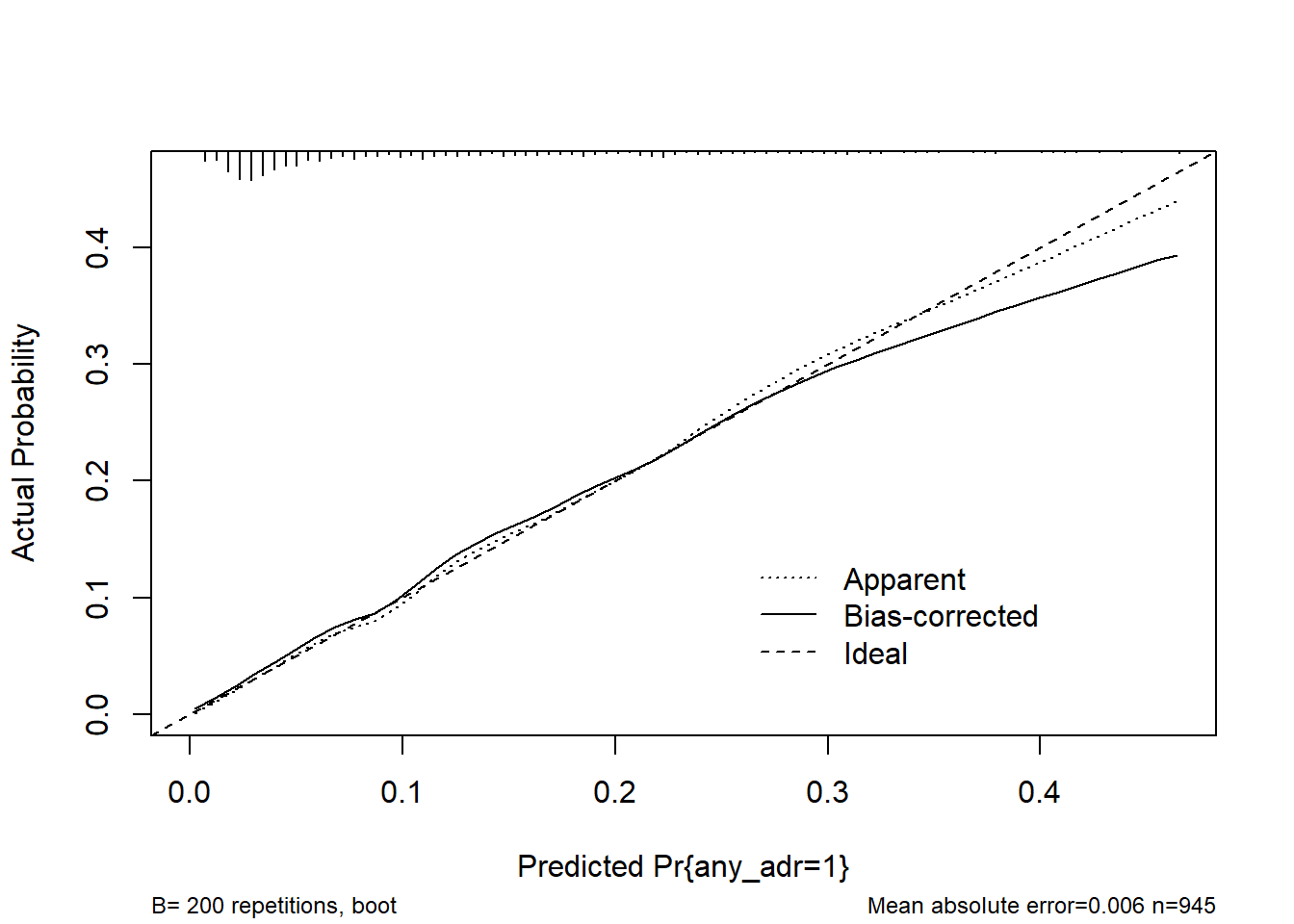

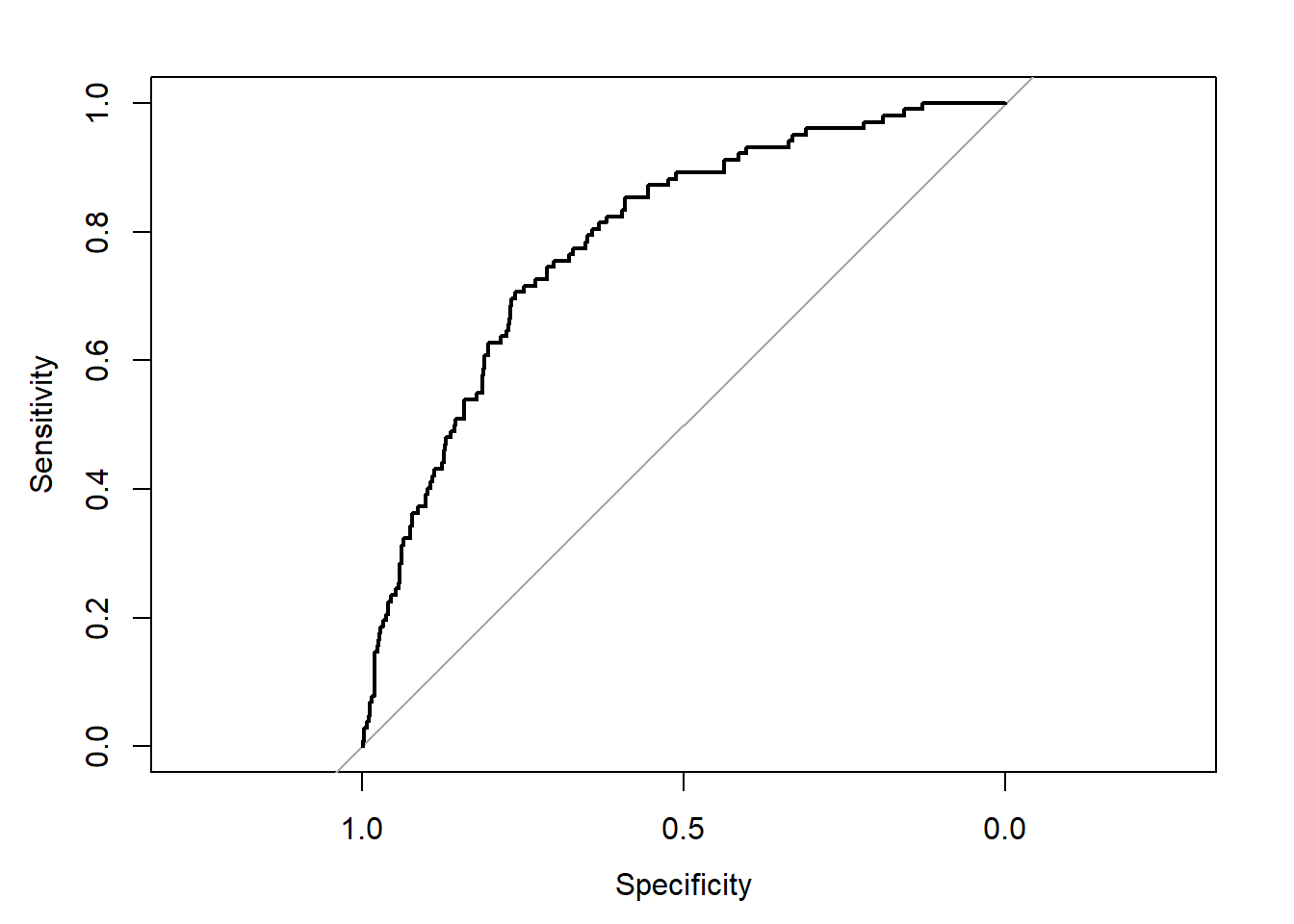

**(A)**

**(B)**

**Footnote**: **(A)** The calibration plot displays agreement between observed and predicted outcome probabilities across deciles of outcome (TB-treatment related adverse drug reaction (TB-ADR)) risk. An ideal calibration curve has an intercept of 0 and a slope of 1 (dashed line). The apparent calibration (dotted line) is calibration of the model in the original data, and the bias-corrected line is corrected for overfitting using 500 bootstrap samples. The bias-corrected calibration intercept and slope were -0.09 and 0.94, respectively. The top of the plot displays a histogram of the distribution of predicted probabilities TB-ADR for the 945 culture-confirmed, drug-susceptible pulmonary tuberculosis participants included in the study. A shrinkage factor of 0.90 was applied to correct uncertainties introduced in model development and improve fit in external validation. **(B)** The receiver operating characteristic (ROC) curve measures discrimination of the model, i.e., how well the model can differentiate between those with and without an outcome. The area under the ROC curve, which is equivalent to the c-statistic, is 0.79 (95% CI: 0.74-0.83).

**Supplemental Figure 3.** The nomogram can be used in clinical settings to estimate individual risk TB treatment-related adverse drug reaction.

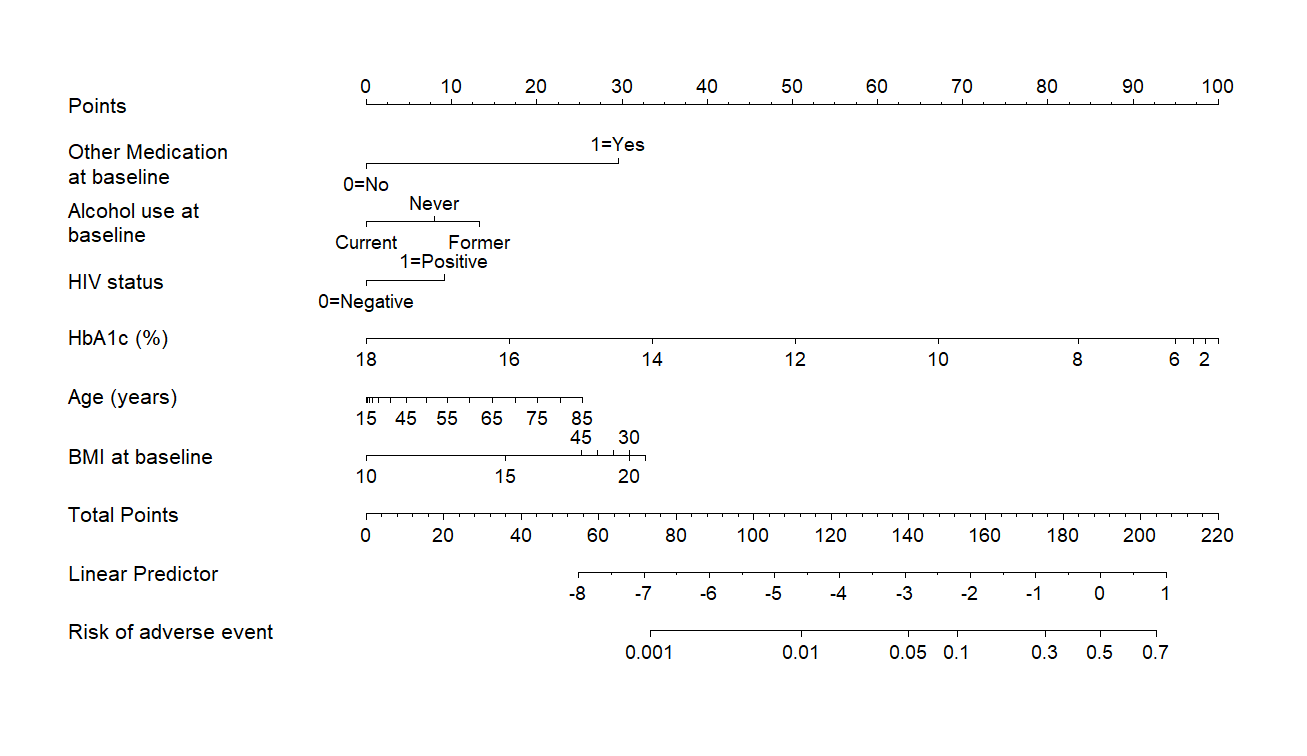

Abbreviations: HIV – human immunodeficiency virus; HbA1c – glycated hemoglobin; BMI – Body Mass Index

**Footnote**: The nomogram can be used in clinical settings to estimate individual risk of TB-treatment related adverse drug reaction. For example, for an individual that has a diagnosis of pulmonary TB and is about to start the treatment. S/he is 55 years old, is using any concomitant medication at baseline (other than the TB treatment), who currently drinks alcohol, is a person living with HIV, has a glycated hemoglobin of 10g/dL, and has a BMI of 20 their risk of developing an adverse drug reaction during the TB treatment can be calculated by: use of other medication at baseline (yes) = 30 points, alcohol use (never) = 0 points, HIV-infection (yes) = 10 points, glycated hemoglobin = 67 points, age = 10 points, BMI = 30 points. Total points = 147, which equates to approximately 7.5% risk of having and adverse drug reaction.

**Supplemental Figure 4.** Adjusted association of HbA1c, BMI, age, and alcohol use with TB-ADR^1^

**(A)**

**(B)**

**(C)**

**(D)**

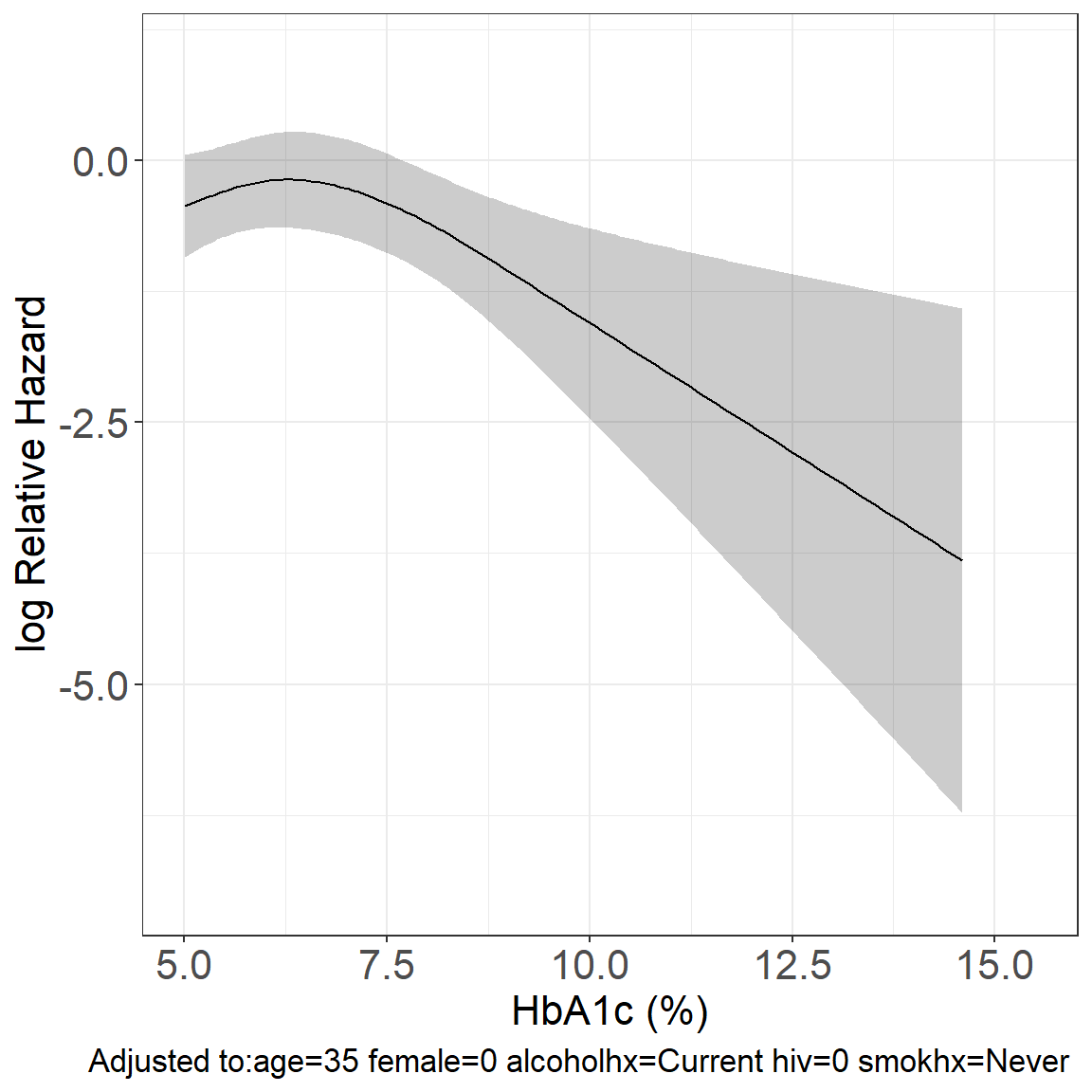

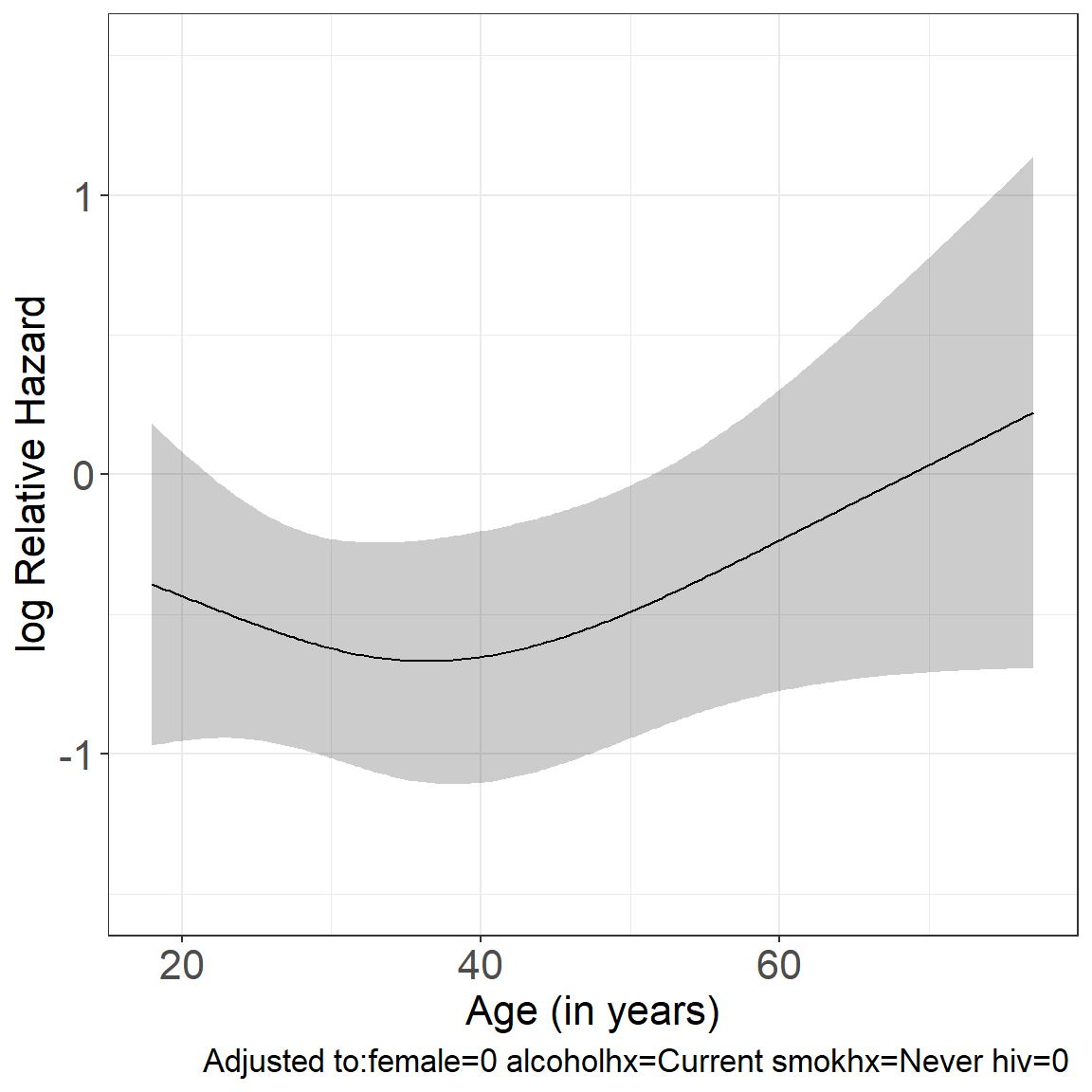

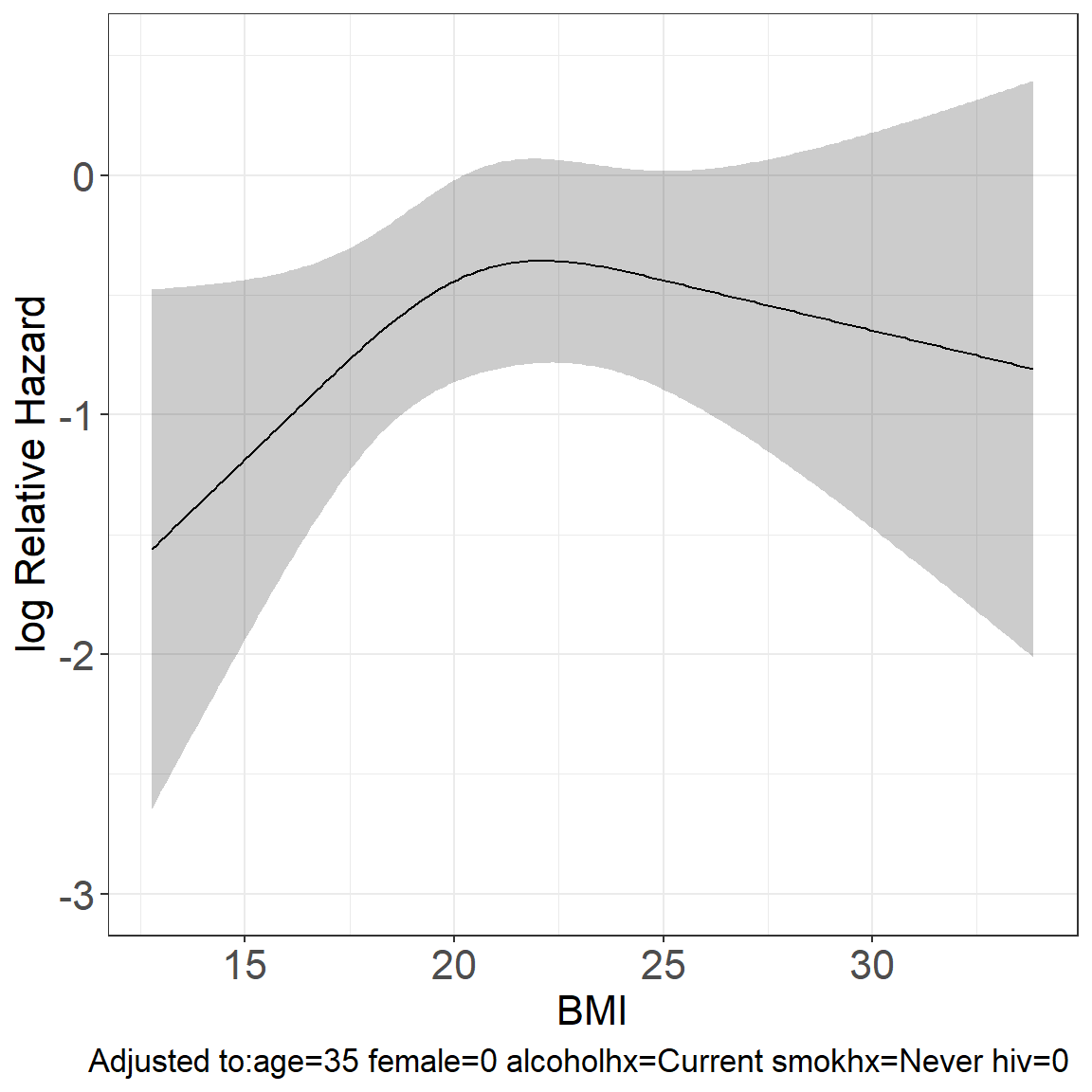

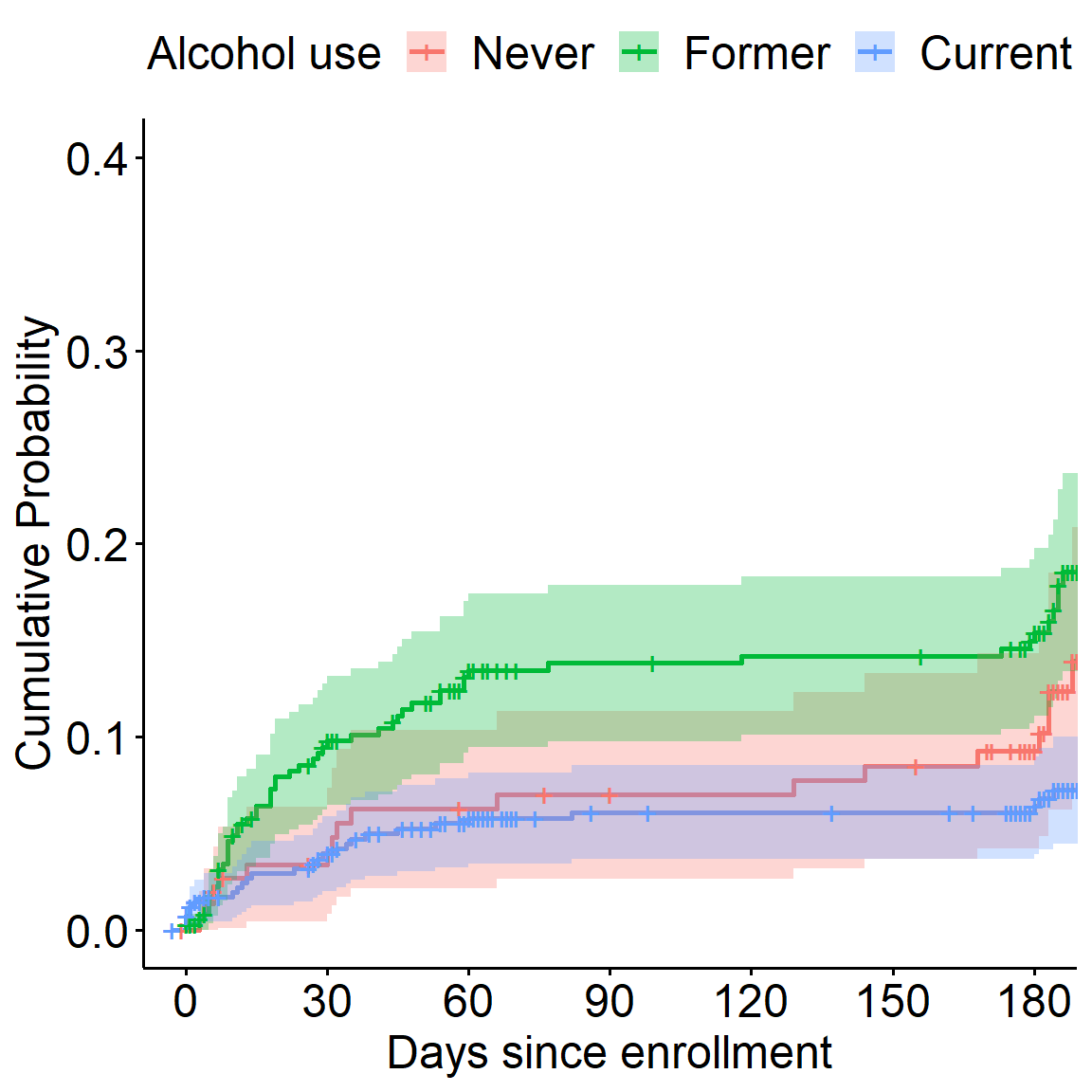

**Footnote:** ^1^ TB treatment-related adverse drug reaction. The log Relative Hazard shown on the plots **(A)** HbA1c, **(B)** Body Mass Index (BMI) are adjusted to an age of 35 years, male sex, and current alcohol use. For **(C)** age (years), adjustment was for sex, alcohol use, and drug use. And for **(D)** alcohol use, adjustment was for sex and age.

**Supplemental Figure 5.** Association between HIV status and HIV-related factors, with ADR.

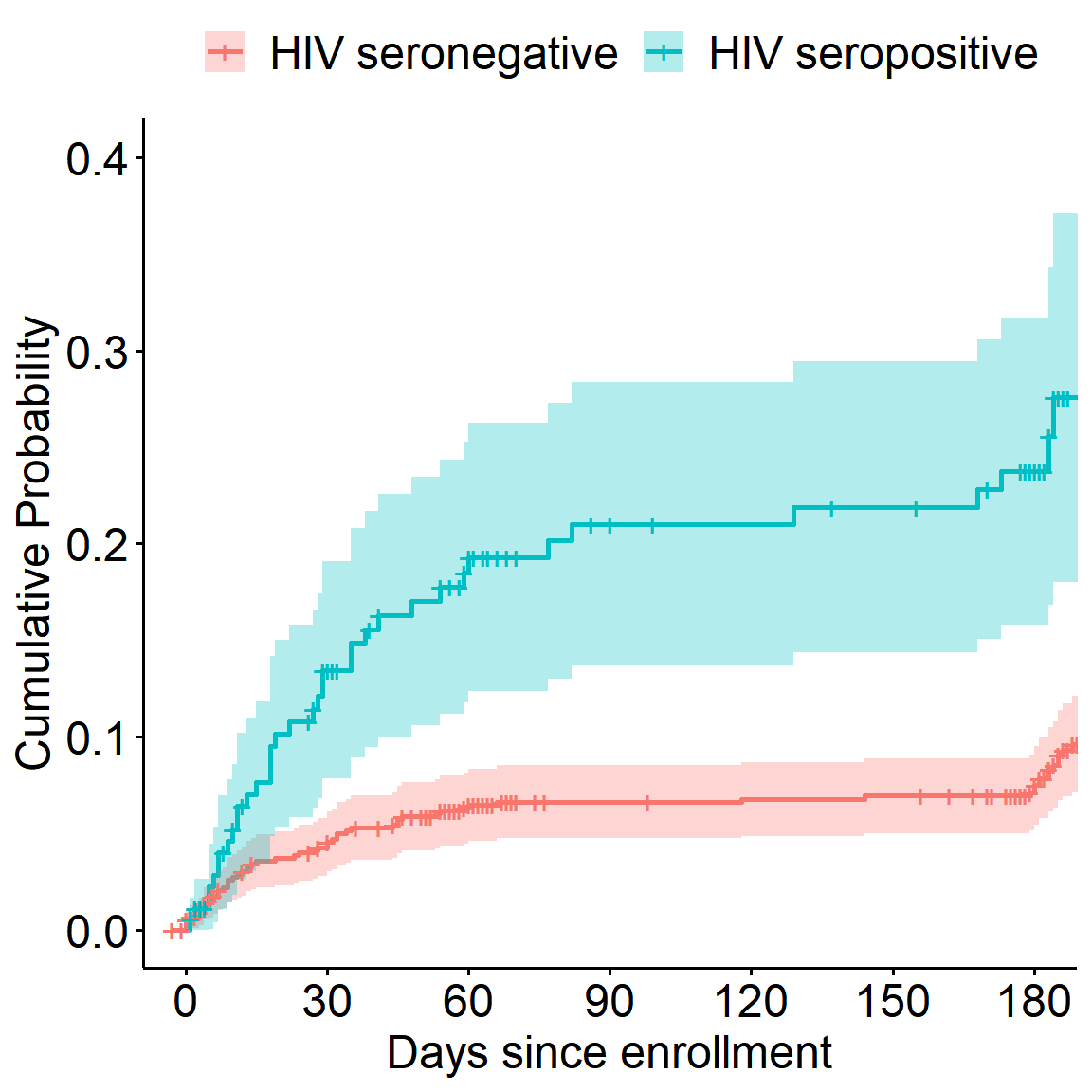

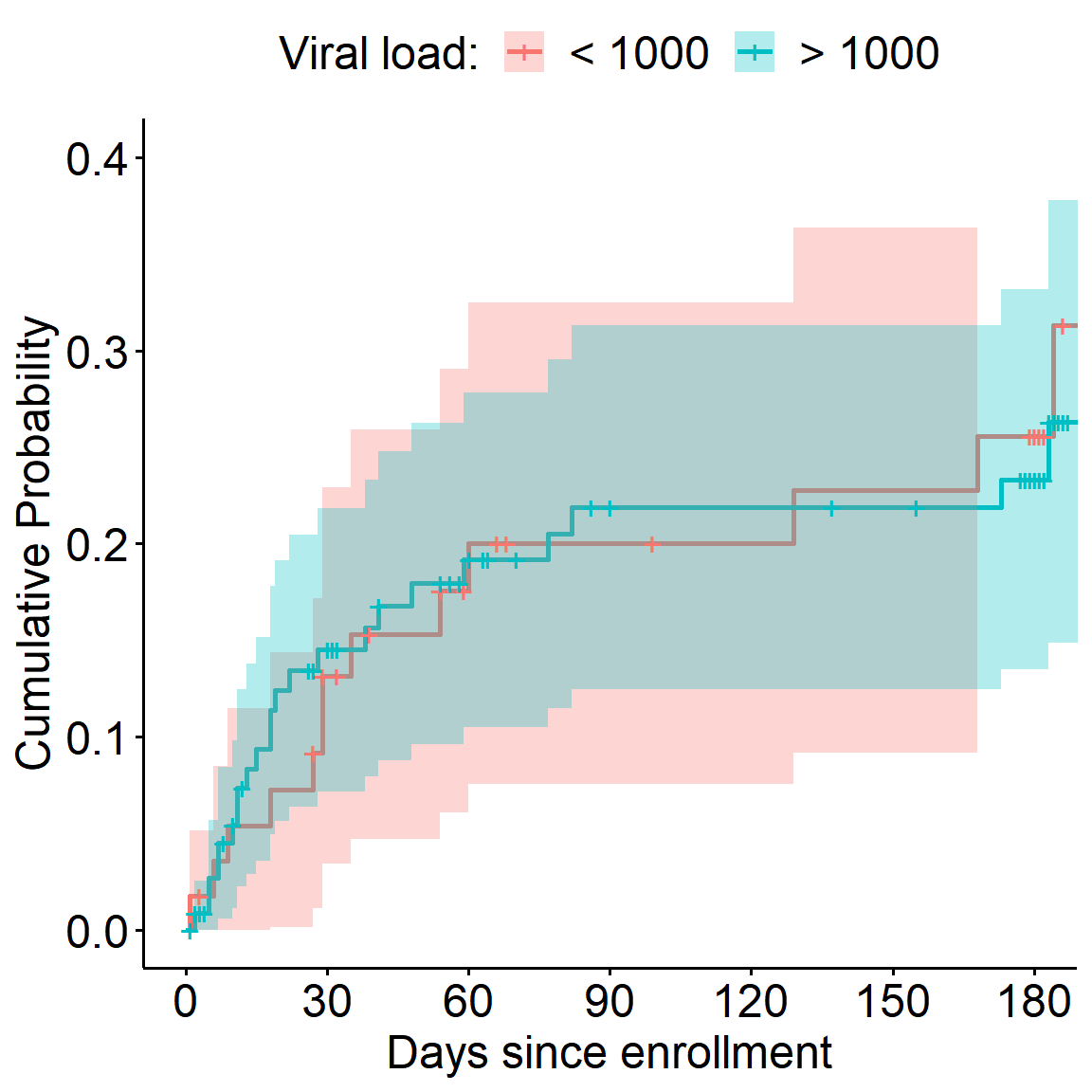

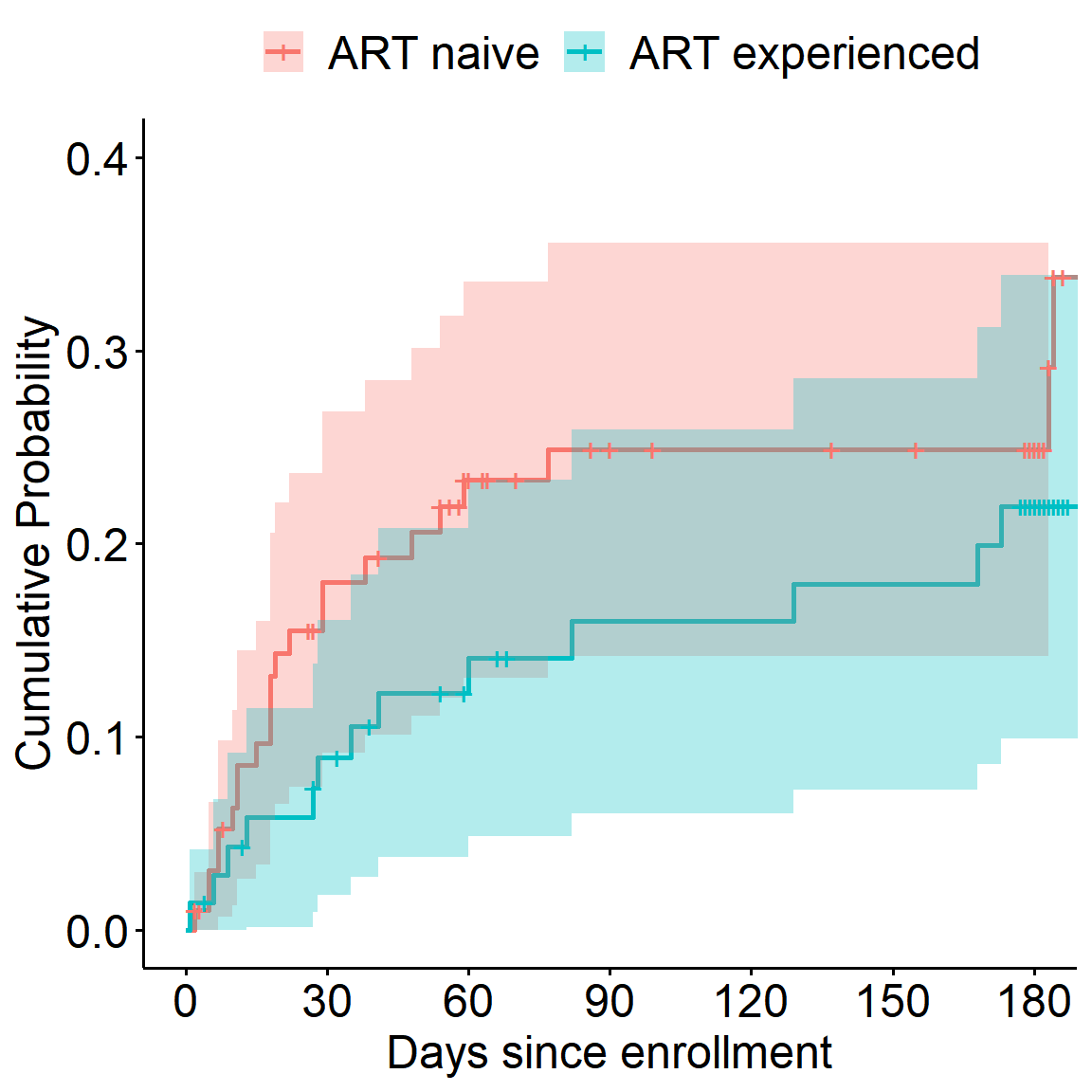

**(A)**

**(B)**

**(C)**

**(D)**

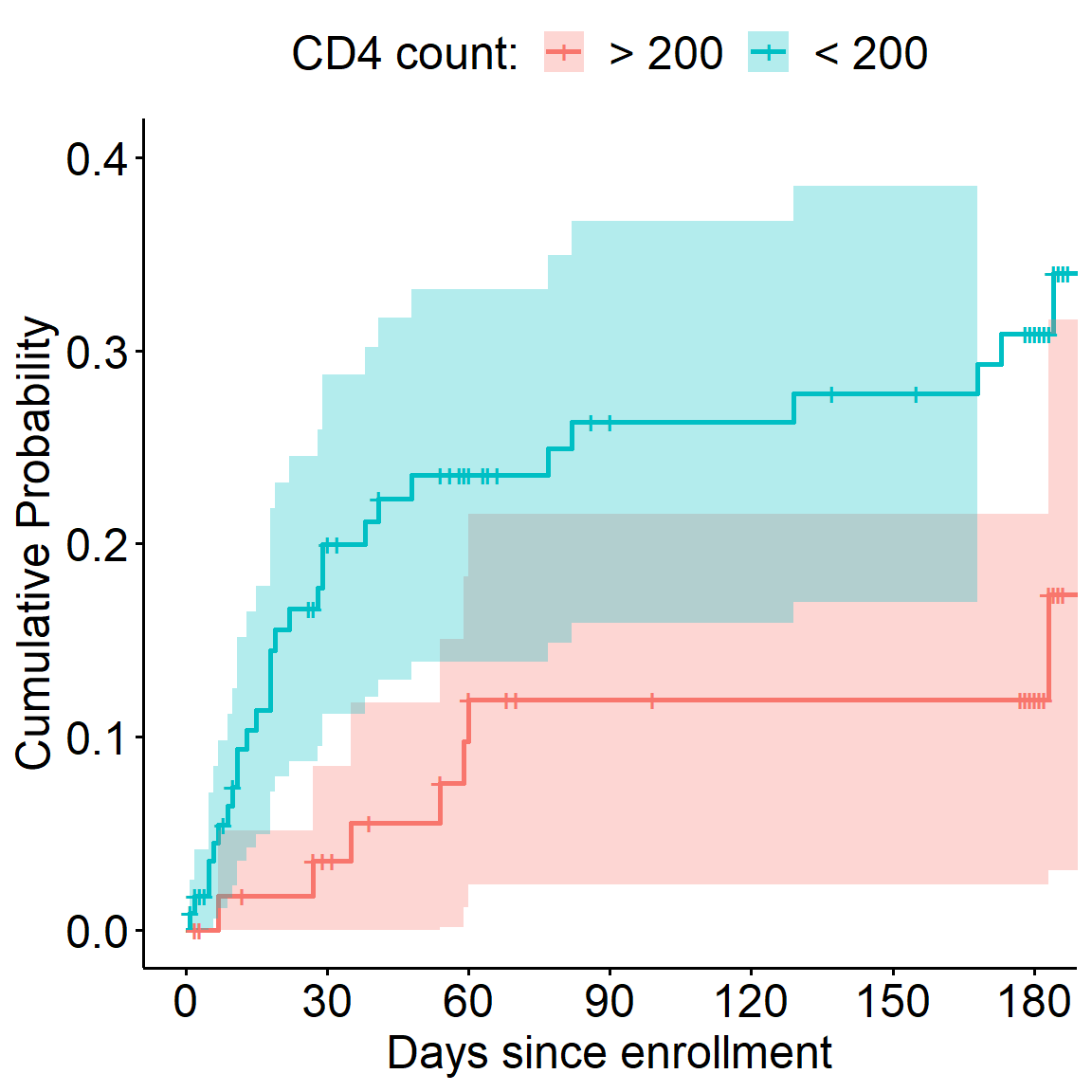

**Footnote:** **(A)** Association of HIV-status with ADR; Association with ADR of **(B)** CD4 cell count (copies/mL), **(C)** viral load (copies/mL), and **(D)** Antiretroviral therapy (ART) status related to TB diagnosis (ART naïve and ART experienced).
